# Radiological phenotyping in COVID-19 Acute Respiratory Distress Syndrome: a secondary analysis of a retrospective cohort study

**DOI:** 10.64898/2026.01.21.26344520

**Authors:** Alexandre Joseph, Jean-Damien Ricard, Constance de Margerie-Mellon, Thaïs Walter

**Affiliations:** APHP, Hôpital Saint-Louis, Department of Anesthesia and Critical Care, Paris, France; AP-HP, Hôpital Louis Mourier, DMU ESPRIT, Service de Médecine Intensive Réanimation, Colombes, France; Université Paris Cité, UMR1137 IAME, INSERM, Paris, France; AP-HP, Hôpital Saint-Louis, Radiology Department, Paris, France; Université Paris Cité, PARCC UMRS 970, INSERM, Paris, France; Université Paris Cité, INSERM U942 MASCOT, Paris, France

**Keywords:** Respiratory Distress Syndrome, COVID-19, Critical Illness, Phenotype, Prone position

## Abstract

**Objective:** To assess whether focal and non-focal COVID-19 ARDS exhibit different respiratory mechanics and arterial blood gas (ABG) trajectories during the first extended prone positioning (PP) session.

**Design:** Post-hoc analysis of a previously published retrospective monocentric cohort study.

**Setting:** A university-affiliated intensive care unit in Paris (France) between March 2020 and April 2021.

**Patients:** Seventy-four adult patients with moderate-to-severe COVID-19 ARDS who underwent extended prone positioning (PP) and had chest imaging (CT or X-ray) performed within five days before the first PP session.

**Interventions:** None

**Measurements:** Changes in compliance, driving pressure, PaO_2_/FiO_2_ ratio, and ventilatory ratio between pre-PP and end-of-PP; chest X-rays and CT scans diagnostic agreement between reviewers assessed by crude agreement and Cohen’s kappa coefficient.

**Main results:** Diffuse ARDS predominated, identified in 91% by the intensivist and 86% by the radiologist. Crude diagnostic agreement was 89% (95% CI 79.8–95.2), while interrater reliability was only fair (κ = 0.44, 95% CI 0.08–0.81). Discrepancies mainly involved focal classifications on chest X-rays, whereas agreement was perfect when CT scans were available. The small number of focal cases precluded comparative analysis of PP trajectories.

**Conclusions:** PP indication or duration should probably not be based on ARDS phenotypes and when differentiating focal from diffuse COVID ARDS is necessary, chest CT scans should be preferred to ensure accurate and reproducible phenotyping.

## Introduction

It has been debated whether COVID-19 related acute respiratory distress syndrome (COVID ARDS) resembles non-COVID ARDS. After adjusting for the timing of intubation during the natural course of ARDS, the prevailing consensus leaned toward the idea that COVID and non-COVID ARDS are similar (1). However, the radiological pattern has been largely overlooked.

Distinguishing focal from diffuse ARDS is the only form of sub-phenotyping associated with personalized therapeutic strategies. The LIVE trial notably demonstrated that focal ARDS may benefit from early prone positioning (PP), without recruitment maneuvers and low PEEP levels, whereas diffuse ARDS may benefit from recruitment maneuvers, higher PEEP levels and no systematic PP (2).

In this study, we have inquired whether focal and non-focal COVID ARDS experience the same mechanistic and arterial blood gas (ABG) trajectories during the first session of extended PP.

## Methods

This study is a post-hoc analysis of a previously published retrospective monocentric study conducted in a university-affiliated ICU. Inclusion and exclusion criteria, as well as the protocol for extended PP, have been described previously(3). Briefly, patients with COVID ARDS were placed in PP when their PaO_2_/FiO_2_ (P/F) ratio was ≤150 mmHg and were maintained in PP for 39-hour PP in median. Chest imaging, either chest X-rays or CT scans, performed within the 5 days preceding the first PP session was reviewed to classify the radiological phenotype. If no CT scan was available, the most recent chest X-ray in the 5 days preceding PP was used for classification.

For all other patients, if a CT scan showed a diffuse pattern at any point within 5 days prior to PP, the patient was classified as diffuse. If the CT scan showed a focal pattern, then two scenarios were distinguished:

- If the CT scan was performed within 48 hours before PP, the patient was classified as focal.
- If the CT scan was performed between 3 and 5 days before PP, the last available chest X-rays before PP was used to evaluate whether the patient had not transitioned to a non-focal phenotype.

A senior ICU physician and an experienced thoracic radiologist independently reviewed all images, blinded to each other’s interpretation. Each imaging set was classified as representing either focal or diffuse ARDS.

Focal ARDS was defined as a loss of aeration strictly limited to a single pulmonary lobe on each side. In contrast, diffuse ARDS was defined as an aeration loss involving two or more pulmonary lobes on at least one side. In chest radiograph interpretation, if an opacity was located over an anatomical zone of lobe overlap, it was assumed by default that only one lobe was involved.

Mechanistic and ABG trajectories during PP were defined as the changes, between pre-PP and the end of PP, in compliance, driving pressure, P/F ratio, and ventilatory ratio.

## Results

A total of 74 patients with imaging available within 5 days prior to their first PP session were analyzed. Of them, 5 (7%) had a CT scan available. Diffuse ARDS remained predominant, identified in 67/74 (91%) of cases by the ICU physician and in 64/74 (86%) by the radiologist. Given this highly unbalanced repartition, no comparisons could be made between trajectories during extended PP in focal and non-focal ARDS.

Crude diagnostic agreement was 89.2% CI95% [79.8; 95.2]. However, the Cohen’s kappa coefficient was 0.44 CI95% [0.08; 0.81], reflecting fair agreement beyond chance. Most discordant classifications involved focal cases, with discrepancies occurring on chest X-rays. In contrast, when chest CT scan data was available prior to PP, diagnostic agreement was perfect.

## Discussion

Our findings indicate that COVID ARDS is predominantly of a non-focal phenotype. Mechanistic and ABG trajectories during 39-hour PP are thus not comparable between focal and non-focal COVID ARDS. Furthermore, as extended PP has been associated with reduced mortality in COVID ARDS, our study does not support the use of ARDS radiological phenotype to guide the indication or duration of PP and thus call into question the LIVE trial protocol (2).

Historically, the main limitation of radiological sub-phenotyping in ARDS has been its poor reproducibility because of the anatomical overlap of the different pulmonary lobes. Our study reaffirms this limitation.

## Conclusion

PP indication or duration should probably not be based on ARDS phenotypes and when differentiating focal from diffuse COVID ARDS is necessary, chest CT scans should be preferred to ensure accurate and reproducible phenotyping.

Further studies are needed to determine not only the most reliable diagnostic strategy for phenotyping ARDS, but also how such phenotyping could impact patient management and outcomes.

## Data Availability

All data produced in the present study are available upon reasonable request to the authors.

## List of abbreviations

COVID ARDS: COVID-19 related acute respiratory distress syndrome
PP: prone positioning
CT: computerized tomography
P/F: PaO2/FiO2
CI: confidence interval
ABG: Arterial blood gas

## Declarations

### Ethics approval and consent to participate

This study was approved by the Ethics Evaluation Committee of Biomedical Research Project (CEERB) Paris Nord (institutional review board—IRB 00006477 of HUPNVS, Université Paris Cité, APHP, reference CER-2021-102) and registered at Clinicaltrials (NCT05124197). Due to the retrospective design of the study, written consent was waived and the registration at Clinicaltrials was completed once we decided to report our experience and results with the extended PP strategy. Patients and families were informed of their right to decline use of their data for the research.

### Availability of data and materials

The datasets generated and/or analysed during the current study are not publicly available due institutional and regulatory restrictions but are available from the corresponding author on reasonable request.

## Acknowledgement and author’s contributions

TW, AJ and CMM collected the data. AJ wrote the first draft. AJ, TW and JDR performed the statistical analysis and contributed to the manuscript. All authors revised and approved the final version. AJ takes responsibility for the integrity of the work, from inception to publication.

